# Environmental Sustainability in England’s NHS Respiratory Prescribing; A grounded theory analysis to define a methodology for estimation of inhaler carbon footprint

**DOI:** 10.1101/2023.05.23.23290379

**Authors:** Helena R.P. Nettleton, Gillian A. Masters, Monica M. Mason

**Affiliations:** Regional Drug and Therapeutics Centre, Newcastle Upon Tyne Hospitals NHS Foundation Trust, UK

## Abstract

**Background:** The impact of climate change on health is increasing, as global warming continues to rise.^1^ Within the health and social care sector, pressurised metered dose inhalers (pMDIs) and breath-actuated pMDIs (BA-pMDIs) have been identified as a significant contributing factor to England’s National Health Service carbon footprint.^2^

**Methods:** A grounded theory study design was applied to formulate a process map and methodology for Inhaler carbon footprint estimation, utilising established inhaler carbon footprint values and pharmaceutical principles.

**Results:** A methodology has been developed to support estimation of inhaler carbon footprint values for those inhalers and refills, that do not have a manufacturer’s independently verified carbon footprint certificate.

**Conclusion:** Definitive Inhaler carbon footprint values, such as those reported here, are required to enable analysis and monitoring of prescribing against Net Zero targets. Further application can also support sustainability assessment and formulary decision making.

## Background: National targets & inhaler carbon footprint

The impact of climate change on health is increasing, as global warming continues to rise.^1^ One strategy to tackle this is to reduce greenhouse gas (GHG) emissions, which will help to both reduce air pollution through reduction in co-emitted particulate matter, and decrease the accumulation of gases, such as carbon dioxide, which absorb and trap heat causing global warming.^2^ NHS England (NHSE) have published a ‘Net Zero’ plan for the National Health Service (NHS) in England, which identifies that pharmaceuticals account for ∼25% of the NHS carbon dioxide equivalent (CO_2_e) emissions (carbon footprint) with pressurised metered dose inhalers (pMDIs) and breath-actuated pMDIs (BA-pMDIs) specifically contributing around 3%.^3^ The NHS Long Term Plan (LTP) sets a target of ‘51% reduction in the carbon footprint by 2025’ for the Health and Social Care Sector, as part of the United Kingdom (UK) Government Climate Change Act (2008).^4^ Furthermore, The NHS LTP outlines that 4% of the total NHS carbon footprint savings are expected to be realised through a ‘shift to lower carbon inhalers’, such as dry powder inhalers (DPIs), equivalent to a 50% reduction in the total inhaler carbon footprint.^4,5^ This would bring the UK in line with Europe and the Scandinavian countries, where pMDI/BA-pMDI use is <50% and 10-30% respectively.^6^

Independently verified manufacturer product carbon footprint (PCF) certificates are currently only available for 39% of the commercially available inhalers marketed in the UK. In order to define the environmental sustainability challenge and monitor progress made against the Net Zero policy and localised respiratory guidance implementation, the inhaler dispensed medicines data (ePACT2) needs to be translated into an actual carbon footprint value. To quantify the locality level Inhaler carbon footprint, each type, strength and pack size of Inhaler needs to be assigned a specific carbon footprint value; estimated as kilograms (kg) CO_2_e, which is then multiplied by the volume of prescribing. Where inhalers have not been formally assessed, the process of estimating a carbon footprint is not an exact science and there are still unknown variables, which may require adjustment for in the future. The method of inhaler categorisation chosen, for example: clinical grouping rather than formulation-based, generalisations such as model of inhaler, and the use of older carbon footprint values; not updated for changes and based on unrevised global warming potential (GWP) values, can have significant impacts on the accuracy of total carbon footprint at the scale of a regional or national level. There is rapid progress in this field both in the number of inhalers that have a carbon footprint certificate and in the number of inhalers which have been re-evaluated and an updated carbon footprint certified. Therefore, it is important to regularly update the data. For instance, the United Nations Environment Programme report (2014)^7^ reported that a DPI typically has a carbon footprint of 7.5- 30 gCO_2_e/actuation, with a midpoint value of 18.75 gCO_2_e/actuation (calculated in 2010). This range and midpoint are currently applied in the NHSE endorsed estimates of DPIs^8^ where the inhaler does not have a manufacturer PCF. However, using updated PCF values for DPIs available in 2022, this study has calculated the mean carbon footprint per DPI actuation to be 10.44 gCO_2_e/actuation (range 3.1 to 25.5 gCO_2_e, SD 6.69, n=32). Pulmicort 400 Turbohaler was excluded due to its carbon footprint being an outlier for a DPI. The difference between the midpoint value and this methodology, when applied to 12 months English prescribing data for DPIs/SMIs and refills is currently 291 Metric tonnes (MT) CO_2_e greater. However, relative to this methodology there is an underestimate of SABA CF of approximately 200 MTCO_2_e. As more pMDI/BA-pMDI prescribing is switched to DPIs and soft mist inhalers (SMIs) and refills are increasingly utilised; shifting the magnitude of prescribing away from pMDI/BA-pMDIs, the variation noted above will become more significant. Undoubtedly though, the best method to estimate the Inhaler carbon footprint impact would be through a change in legislation, requiring all pharmaceutical companies to assess and report on the carbon footprint of their products.

In the absence of this, several review publications have reported on inhaler carbon footprints.^9-12^ In a key paper utilised to underpin the NHSE endorsed values, Wilkinson et al., (2019) published an economic and footprint analysis for the UK NHS inhaler prescribing, which used a number of sources to estimate the range and mid-point inhaler carbon footprint values when categorised by therapeutic class.^9^ There were limitations to this research, which included:

i. Grouping inhalers by therapeutic type. This approach does not consider formulation variables such as the Active Pharmaceutical Ingredient (API) and the presence of co-solvents, which affect the volume of propellant required.^13^
ii. Assigning an estimated value to inhalers which have a PCF certificate.
iii. Comparing full life cycle PCF values with estimated ‘Use’ phase values.
iv. Assigning an estimated value of 1 KgCO_2_e to all DPIs.
v. Not considering the use of refills.

In an attempt to address the above limitations, this study used a grounded theory approach to explore the hypothesis that there are key features in the formulation specifics of inhalers that would enable categorisation of, and improve the accuracy of, estimating inhaler carbon footprint values. Birks and Mills (2015) described grounded theory methods as ‘inductive in that they are a process of building theory up from the data itself. Induction of theory is achieved through successive comparative analyses.’^14^

The researchers collated the available manufacturer PCF data, primary research papers and inhaler formulation specifics. These data were then used to generate a set of assumptions which enables carbon footprint values to be estimated for all currently available inhalers. The Inhaler carbon footprint values, when applied in the Regional Drug and Therapeutics Centre (RDTC) Inhaler Carbon Impact Assessment Tool^15^, has facilitated an increased understanding of health service sustainability in prescribing performance and has been used to support commissioning level decisions regarding the financial and carbon impact of inhaler prescribing.

## Methods

All pharmaceutical manufacturers were contacted and asked to provide carbon footprint data for all respiratory inhalers that they manufacture or hold the product license for. Where an independently verified manufacturer’s carbon footprint was provided, this value was assigned to the specific inhaler and strength to which it applied. There is currently PCF data for only 39% of the 143 inhalers licensed in the UK (updated in November 2022). The pharmaceutical company response rate to requests for PCF information was 64%, with only 41% of the companies able to provide actual PCF values. To estimate carbon footprint values for the remaining devices, a literature search was carried out with the purpose of identifying specifics about the inhalers, including propellant type and volume, ethanol content, canister weight and APIs; where not provided by the manufacturers. Sources included: NICE Healthcare Database Advanced Search, British National Formulary, American Society of Health System Pharmacists (AHFS) Drug Information and Summaries of product characteristics (accessed via the electronic medicines compendium).

A number of carefully constructed assumptions and calculations were then made, using a grounded theory study design ^16^ (Figure 1).

**Figure 1.**
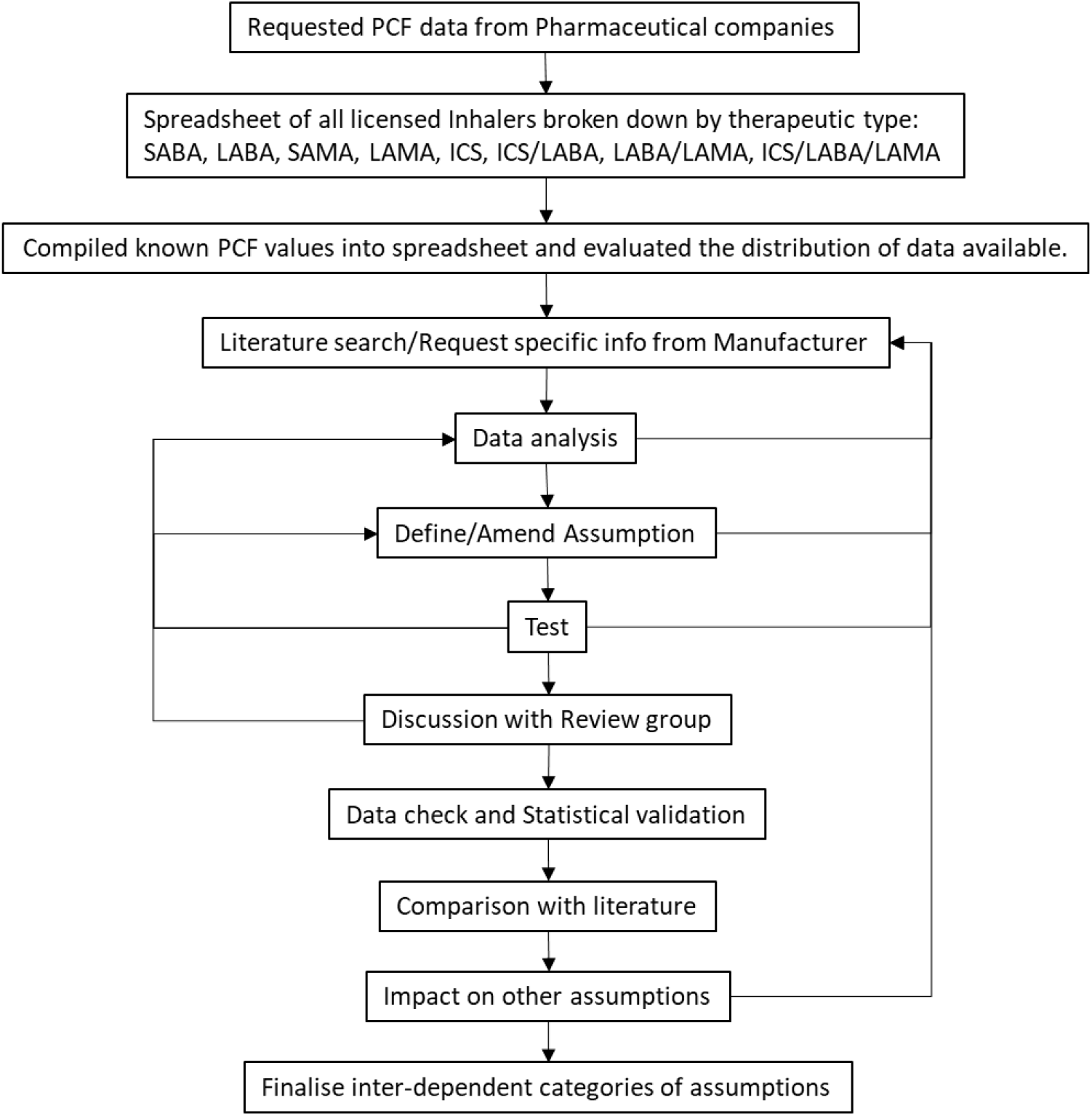
Grounded theory approach applied to quantitative data study method to define categories of assumptions required to estimate Inhaler carbon footprints.

This method utilises the available quantitative data to enable comparative analysis and identification of appropriate categories based on inhaler formulation characteristics, rather than based on the therapeutic classification of the drug. In the test stage, multiple analytical methods were applied where relevant to narrow down, exclude and refine the use of scientific literature that referred to formulation specifics of different inhalers. Calculations utilising different datasets and cross referencing with known carbon footprint values were similarly used to refine the accuracy of estimation methods and exclude relationships. For example, using the mL/actuation value determined experimentally^17^ alongside Avogadro’s gas constant, it was possible to estimate the weight of propellant in a Clenil Modulite pMDI. This estimated propellant weight, in conjunction with the constants and assumptions applied in this study, returned an overall estimate of Inhaler carbon footprint of 15.96 KgCO_2_e/inhaler versus 16.38 KgCO_2_e/inhaler (average of the four PCF values for the different strengths of Clenil Modulite pMDI). The methodology reported here was considered by the researchers to be a more accurate model than using midpoints by therapeutic class, which returned an estimate of 20.35 KgCO_2_e/inhaler.^9^ When the two methods were multiplied up by the volume of prescribing for pMDIs/BA-pMDIs in England over 12 months, the difference in carbon footprint (excluding inhalers with a manufacturer provided value) using this methodology was found to be as follows: Inhaled corticosteroids (ICS) -6,100 MT CO_2_e, ICS/LABA +4,534 MTCO_2_e and LABAs +714 MTCO_2_e. The calculations from this methodology have been updated to use the latest GWP values (2022), whereas the midpoint estimates are based on the inhalers which had PCFs calculated prior to 2019, using lower GWP values. There was a negligible difference between SABA, SAMA and LAMA pMDI/BA-pMDI total CF as most of these inhalers now have PCF values. Although overall the difference is minimal, as policy decisions drive changes in England towards a greater proportion of DPI/SMI prescribing and the use of refills, in addition to increased prescribing of ICSs and the current problematic overuse of SABAs, the methodology currently endorsed may represent a significant underestimate of carbon impact.

The review group, made up of two senior pharmacists and a statistician, were tasked throughout the study with evaluating the ongoing narrative, method and rationale of principles & comparative analytical methods applied to ensure a robust systematic process. Primary data was discussed and statistical power and significance considered. The method required a cyclical approach enabling adaptation of categories as more data becomes available and subsequently identified the opportunities for using other additional data to inform calculations. The assumptions and constants shown here are the result of the fourth update of the data in November 2022 (6 monthly updates scheduled) and re-definition of the categories required through comparative analysis. These ongoing systems ensure that the methods and processes remain valid in this rapidly developing field. A final peer review of the overall data, categorisations, method and inter-relationships was conducted by senior pharmacists and medicines information & evaluation scientists. All manufacturers were contacted to advise them of the outcome of this process with respect to their inhalers only and the assumptions made for their inhalers (if required). They were offered the opportunity to comment and provide further relevant information.

## Results

### Inhaler carbon footprint full life cycle value estimation

A number of factors have a significant impact on the inhaler carbon footprint value and have been accounted for in this analysis. These include propellant type and volume, ethanol concentration, overage of doses, type of API, inhaler device type and refills. The categories defined through this study alongside the method, constants, equations and reference products defined to enable a more accurate estimation of Inhaler carbon footprint, are summarised in Figure 2.

**Figure 2.**
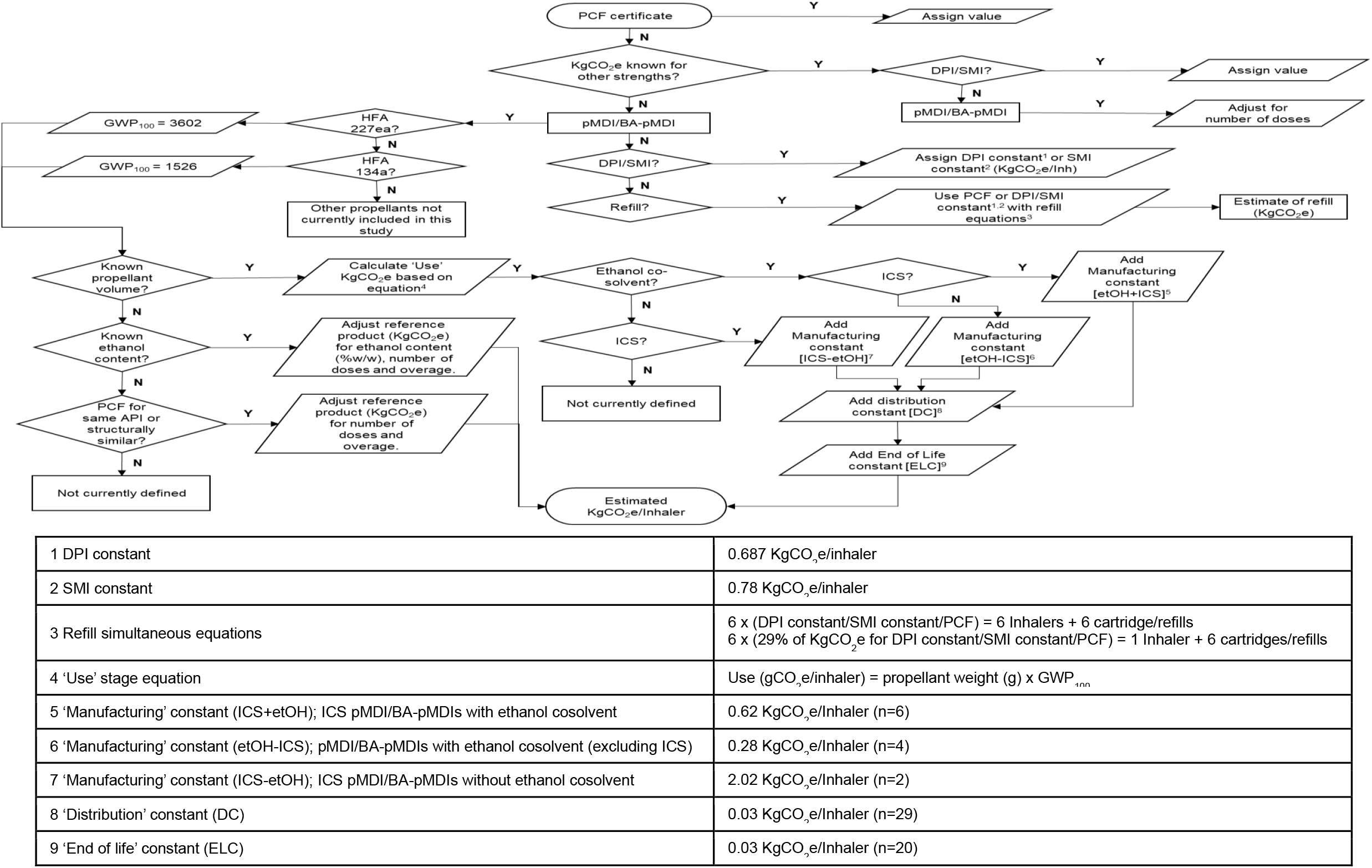
Process map of the methodology for the estimation of pMDI/BA-pMDI, DPI or SMI carbon footprint.

Manufacturer provided PCFs consider the whole lifecycle of an inhaler, with 4 main phases:

- Manufacture
- Distribution
- Use
- End of life

Full life cycle analysis should compare like with like as far as possible. Where carbon footprint data was reported in further subdivisions, it has been merged to present values in a standardised manner representing the four main phases outlined above.

#### pMDIs/BA-pMDIs

Both pMDIs and BA-pMDIs use propellants to deliver the active substance, and the global warming potential (GWP) of these propellants varies. The RDTC applied the following values for GWP over a 100 year timescale (GWP_100_) for hydrofluoroalkane (HFA) propellants:

- HFA 134a = 1526
- HFA 227ea = 3602

These values correspond with those quoted in the data tables by the Intergovernmental Panel on Climate Change.^18^ It is important to state the reference values used in any calculation as there is variability in those quoted in the literature.^9,19,20^

The manufacturer reported carbon footprint values for pMDI/BA-pMDIs containing the propellant HFA 134a, typically range from 11.24 to 28.26 KgCO_2_e/inhaler (n=18), with an average of 15.83 KgCO_2_e/inhaler (SD=3.95). The solubility of the APIs varies with chemical structure, requiring different quantities of ethanol (co-solvent) and propellant. ^13^ This is reflected in the broad range of inhaler carbon footprint values and the large SD noted above. Variation is also seen within therapeutic class, when there are enough carbon footprint values reported to enable comparison. For instance, Inhaled corticosteroid/Long-acting beta-agonist (ICS/LABA) inhalers with the propellant HFA 134a range from 11.25 to 19.29 KgCO_2_e/inhaler with an average of 14.66 KgCO2e/inhaler (n=5, SD=2.9). As such, the use of range values for categorisation by inhaler therapeutic class were not applied in this work, as they represent a broad distribution and do not account for the basic differences in pharmaceutical formulation.

To ensure a uniform dose is produced each time the pMDI/BA-pMDI is actuated, inhalers contain an excess of drug and propellant, typically called “overage”.^17^ Any carbon footprint estimations should include the entire amount of propellant per whole device. Where manufacturers’ report a carbon footprint per actuation value then the number of ‘additional doses’ or ‘overage’ in an inhaler is also required, as this will include additional propellant. However, clarification needs to be sought from the manufacturer to confirm whether the overage is included in the calculation of ‘carbon footprint per actuation’. Technically a misrepresentation by definition but there are examples of this. For the purpose of this work, when a manufacturer has not provided information on overage, a conservative estimate of 10% was calculated to reflect the overage of ‘complete doses’. This is a minimum value that should be applied and is consistent with the literature^17^ and commercial in confidence data provided by manufacturers regarding their formulation specifics.

Wilkinson et al (2019) proposed that the majority of the carbon footprint of a pMDI/BA- pMDI is attributable to the propellant content and that it could be estimated using the formula: propellant weight (g) x GWP_100_ = gCO2e/inhaler.^9^ The authors used the canister weight to infer the propellant weight as this is the major constituent. However, this does not adjust for the inclusion of ethanol as a co-solvent, which reduces the amount of propellant needed.^13^ It also does not consider other phases of the carbon footprint life cycle. Therefore, this assumption was only applied in the RDTC analysis as a ‘Use’ phase, in conjunction with an adjustment in the calculations equivalent to the ethanol content present (detailed below).

Where the value for the ‘Use’ phase of a pMDI/BA-pMDI life cycle has been calculated from propellant weight (manufacturer provided or value taken from the literature), the appropriate RDTC calculated constant for ‘manufacturing’, ‘distribution’ and ‘end of life’ phases have also been included in the overall value determined to reflect a whole lifecycle. There are different RDTC constants calculated for each phase for pMDIs/BA- pMDIs, depending firstly on whether they contain ethanol and then differentiated by type of API (ICS or non-ICS). The RDTC calculated constants (Figure 2) are based on all the data provided from the inhaler manufacturers and as such are evolving as more data becomes available.

From the collated manufacturer provided data, it is possible to evaluate the impact of the category variables. For instance, the carbon footprint for the manufacturing phase is affected by factors including the APIs and solvents.^13^ When an ICS was one of the APIs in an ethanol containing pMDI, the average manufacturing carbon footprint increased by 62% compared to a non-ICS API.

In light of the above, when there is no known information on the specific formulation of an inhaler, it is considered appropriate to estimate the carbon footprint based on a ‘reference product’. Selected reference products were matched for API (or structurally similar if identical API not available), inhaler type and presence or absence of ethanol. For example, Clenil Modulite was used as an ICS reference product for carbon footprint, using an average of the 4 manufacturer provided values. Clenil Modulite has a known ethanol content of 15% w/w^21^; which is within the range of the optimum concentration (10-15% w/w) required for maximum respirable mass of beclometasone in aerosol formulations.^22^ To estimate the carbon footprint of a beclometasone pMDI/BA-pMDIs, the known ethanol content of the inhaler to be estimated was used to calculate the maximum propellant volume present and make a proportional change in the known carbon footprint of the reference product. Similarly, Serevent 25 micrograms pMDI was used as a LABA reference product for carbon footprint (manufacturer provided) and has a known ethanol content of 0% w/w^23^, and therefore an assumed propellant volume of 100%. Factors such as the number of doses per inhaler were also accounted for.

#### Dry powder inhalers

DPI manufacturer provided carbon footprint values range from 0.282 to 1.63 KgCO_2_e/inhaler, but are typically less than 1 KgCO_2_e with the exception of some strengths of Turbohaler (n=32).

However, it is considered here that the carbon footprint value per actuation should not be used to estimate DPI carbon footprints by multiplying by the number of actuations in an inhaler, as the relationship between DPI carbon footprint and number of actuations is not linear and has a very weak correlation (r^2^=0.088, correlation coefficient = 0.4004). Only a proportion of the DPI carbon footprint is attributable to the APIs and would thus be affected by increasing the number of doses. For instance, Ellipta^24^, Accuhaler^25^ and Breezhaler^26^ devices report that 0.5 to 2%, 1 to 29% and 15.8 to 22.5% respectively of their carbon contribution is due to the APIs.

The impact of variables such as API and specific DPI type are indistinguishable within the dataset available. Therefore, due to the relatively narrow range of DPI carbon footprint values and the relatively large sample size, an average DPI carbon footprint value per device was calculated from a sample including the following DPI types; Ellipta, Turbohaler, Accuhaler, Breezhaler, Easyhaler, Handihaler and NEXThaler. This average DPI carbon footprint value has been applied in RDTC analysis as an estimate for all DPIs that do not have a manufacturer provided carbon footprint certificate.

#### Soft mist inhalers

SMIs are distinctly different in design from DPIs, so a separate average SMI carbon footprint has been calculated. There was only a negligible difference in the contribution due to the different APIs; 0.465 KgCO_2_e for ipratropium & Fenoterol (not available in the UK) versus 0.456 KgCO_2_e/inhaler for tiotropium.^11^ Although there is currently only a sample size of 2 manufacturer provided carbon footprint values, the variation across the range is small at 0.009 KgCO_2_e.

#### Refills

Refillable inhalers have 2 dispensing components identifiable in the prescribing data; the device & refill cartridge/capsules combined and the refill cartridge/capsules alone. In order to accurately determine changes in total carbon footprint, the values assigned need to be reflective of the different combinations. However, because the refillable inhaler is re-usable, the carbon footprint value is directly related to the refill value and changes with the time period over which the inhaler is used. For instance, an inhaler & refill combination used monthly has a different footprint to an inhaler & refill used in month 1 followed by 5 months of refills, which continue to utilise the inhaler from month 1. The ‘Use’ phase footprint of the inhaler in the second scenario will be different and so they cannot simply be considered as independent. Hansel et al., (2019) measured a reduction in product carbon footprint of 57% and 71% at 3 months and 6 months respectively, when refills were used.^11^ The percentage reduction in combination with simultaneous equations (below) have been applied here to estimate a refill carbon footprint for both DPIs and SMIs, assuming Inhaler re-use and use of refills over six months. The time-period of six months was chosen to reflect the data in the literature and licensing for currently available refillable DPI/SMIs.

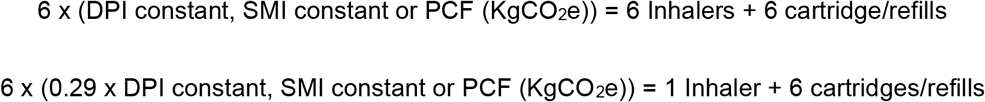

#### Alternate strength of inhalers

Typically, manufacturers have provided carbon footprint values for a single strength of an inhaler, as it is generally considered that the contribution of the API concentration has a negligible impact on the total carbon footprint. Where manufacturers have not provided data for all strengths of an inhaler, the value provided (or average of values) has been applied to the alternate strengths.

Seven examples of carbon footprint data for multiple strengths of the same inhaler have been evaluated here. DPIs showed a variation of 0 to 0.47 KgCO_2_e/inhaler in total carbon footprint across the different strengths of API for the same inhaler (n=12), whereas pMDIs showed a greater variation of 0.46 to 2.9 KgCO_2_e per inhaler (n=6). It is worth noting that all the pMDIs evaluated contained an ICS. The carbon footprint values of pMDI/BA-pMDIs were not available for multiple strengths of other APIs. All the inhaler ICSs are glucocorticoids and therefore have hydroxyl acids.^27^ The manufacturers stated that fluticasone is “a highly carbon intensive API input due to the presence of hydroxyl acids, compared to other APIs and the higher product strength of products based on this API”.^25^ Therefore, inhalers with an ICS as the API will have a higher relative carbon footprint, which will increase with higher strengths of ICS formulation. All manufacturer provided values were for the highest strength of that inhaler, therefore potentially resulting in an overestimate of the footprint for lower strength inhalers. The exception to the above point was for four DPIs, where the carbon footprint provided by the manufacturer was not for the highest strength formulation. However, the variation noted above for DPIs is less significant than for pMDIs and so the alternate strength values were applied in this study carrying this potential error.

The alternative of using a ‘class average’ for the ICS DPIs is considered to be less accurate due to the variation in both the ICS as well as strength, which would have yielded higher carbon footprints, For example:

##### ICS DPI

- 1.02 KgCO_2_e/inhaler (ICS DPI average) versus 0.65 KgCO2e/inhaler (lower strength of same DPI) and compared to the overall DPI average of 0.687 KgCO_2_e/inhaler.

##### ICS/LABA DPI

- 0.715 KgCO_2_e/inhaler (ICS/LABA DPI average) versus 0.48447 KgCO_2_e/inhaler (lower strength of same DPI) and compared to the overall DPI average of 0.687 KgCO_2_e/inhaler.

## Conclusion

Estimation of inhaler carbon footprints is complex and relies on estimation of, and adjustment for, a number of factors. The methodology discussed above has been used to produce the RDTC’s Inhaler Carbon Impact Assessment Tool, which is regularly reviewed and updated to ensure it remains robust.

The Regional Drug and Therapeutics Centre is an NHS funded organisation in England. We are entirely independent of the pharmaceutical industry and do not receive any funding from them. There are no conflicts of interest to declare.

## Data Availability

All data produced in the present study are available upon reasonable request to the authors

## Notes

### Competing Interest Statement

The authors have declared no competing interest.

### Funding Statement

This study did not receive specific funding, resource was allocated to this work stream through current funding from multiple primary care stakeholder CCGs.

